# Genetic characterization of Parkinson’s Disease in a Chilean cohort

**DOI:** 10.64898/2026.08.04.26358901

**Authors:** Paula Saffie-Awad, Peter Wild Crea, Spencer Grant, Paul Suhwan Lee, Thiago Peixoto Leal, Daniel Teixeira-dos-Santos, Fulya Akçimen, Marzieh Khani, Emily Waldo, Ximena Pizarro-Correa, Ernesto Solís-Añez, Cornelis Blauwendraat, Andrew Singleton, Christine Klein, Sara Bandres-Ciga, Ignacio Fernandez Mata, Miguel Inca-Martínez, Artur F. Schumacher Schuh, Pedro Chaná-Cuevas

## Abstract

The genetics of Parkinson’s disease (PD) in underrepresented populations remain poorly characterized, potentially overlooking population-specific contributions. We analyzed 461 Chilean PD cases from a movement disorders center. Pathogenic, likely pathogenic, or *GBA1* risk variants were identified in 58 individuals (12.6%), mainly in *LRRK2* (50%) and *GBA1* (44.8%), while *PRKN, SNCA*, and *SQSTM1* collectively represented 5.2%. All *LRRK2* variants were p.G2019S, with an overall frequency of 6.3%, the highest reported in South America, and enrichment of Ashkenazi Jewish ancestry at this locus. These findings characterize the genetic landscape of PD in Chile and support its relevance for *LRRK2*-targeted studies.

## INTRODUCTION

Parkinson’s disease (PD) results from the interplay of genetic and environmental factors, with genetics playing a major role in familial and early-onset cases [1, 2]. Approximately 10-15% of patients worldwide carry an identifiable pathogenic or risk variant [3, 4], which is increasingly relevant for prognosis, genetic counselling, and eligibility for gene-specific clinical trials and emerging targeted therapies [5]. However, PD genetic research is largely derived from European-ancestry cohorts, and studies from South America account for only ~10% of those conducted outside Europe and North America [6], thereby limiting variant interpretation, risk estimation, and generalizability across populations [7, 8].

South American populations are shaped by varying proportions of African, European, and Native American ancestry, resulting in substantial admixture and population structure that can contribute to differences in allele frequencies and variant distributions [9]. Chile illustrates this regional admixture profile, with ancestry estimates of approximately 52% European, 45% Indigenous (mainly Mapuche and Aymara), and 3% African [10], allowing assessment of population-specific variants, including patterns consistent with founder effects. Local studies remain scarce, with fewer than 200 patients analyzed across seven reports [11]. The largest study (n=166) identified the *LRRK2* p.G2019S variant in approximately 3% of cases, consistent with other South American cohorts [12]. Few studies have examined additional PD genes in Chilean patients [13] and clinical–genetic correlations and ancestry-related patterns remain poorly characterized.

To address these gaps, we aimed to define the frequency and clinical correlation of genetic forms of PD in Chile, including *GBA1* as a major risk factor. This work provides a reference framework for region-specific diagnostic strategies and future gene-informed research in Chile and the broader region.

## MATERIALS AND METHODS

We recruited PD patients at the Chilean site of the Latin American Research Consortium on the Genetics of Parkinson’s Disease (LARGE-PD) based at a movement disorders center in Santiago, Chile. All patients who fulfilled the United Kingdom Parkinson’s Disease Society Brain Bank criteria for PD [14], were eligible, regardless of age at onset (AAO) or family history (FH) of PD. Clinical data were collected by a movement disorders specialist using the standardized LARGE-PD questionnaire [15]. Motor and non-motor variables were recorded as clinician-reported items predefined in the instrument (Table 1) and a cognitive screening was performed using the Montreal Cognitive Assessment. The study was approved by the research ethics committee, and all participants provided written informed consent.

**Table 1.** Demographic and clinical characteristics of the cohort.

| Variable | All (n=461) | Positive (n=58) | Negative (n=403) | p-value |
| --- | --- | --- | --- | --- |
| Age at baseline | 63.40 ± 11.78 | 62.00 ± 10.47 | 63.61 ± 11.95 | 0.287 |
| Age of onset | 55.51 ± 12.17 | 53.84 ± 11.11 | 55.75 ± 12.31 | 0.233 |
| Time to diagnosis (years) | 1.54 ± 2.46 | 1.71 ± 3.03 | 1.51 ± 2.37 | 0.643 |
| Education (years) | 13.56 ± 4.12 | 14.21 ± 3.16 | 13.47 ± 4.24 | 0.115 |
| Disease's duration (years) | 7.96 ± 6.23 | 8.16 ± 5.82 | 7.94 ± 6.29 | 0.791 |
| Early onset (≤50 years) | 154 (33.4%) | 22 (37.9%) | 132 (32.8%) | 0.458 |
| Male | 272 (59.0%) | 34 (58.6%) | 238 (59.1%) | 1.000 |
| Family history | 156 (33.8%) | 31 (53.4%) | 125 (31.0%) | **0.001** |
| Rest tremor | 340 (73.8%) | 45 (77.6%) | 295 (73.2%) | 0.527 |
| Hyposmia | 207 (44.9%) | 31 (53.4%) | 176 (43.7%) | 0.204 |
| Constipation | 198 (43.0%) | 21 (36.2%) | 177 (43.9%) | 0.321 |
| Depression | 190 (41.2%) | 27 (46.6%) | 163 (40.4%) | 0.395 |
| Anxiety | 206 (44.7%) | 30 (51.7%) | 176 (43.7%) | 0.261 |
| Sleep problems | 227 (49.2%) | 35 (60.3%) | 192 (47.6%) | 0.091 |
| RBD | 171 (37.1%) | 20 (34.5%) | 151 (37.5%) | 0.772 |
| Levodopa response | 352 (76.4%) | 44 (75.9%) | 308 (76.4%) | 1.000 |
| Levodopa induced dyskinesias | 121 (26.2%) | 22 (37.9%) | 99 (24.6%) | **0.038** |
| MOCA ≤25 | 268 (58.1%) | 28 (48.3%) | 240 (59.6%) | 0.118 |
| H&Y stage ≤2.5 | 331 (71.8%) | 36 (62.1%) | 295 (73.2%) | 0.087 |

Characteristics are shown for the entire cohort and stratified by genetically positive and genetically negative status. Continuous variables are presented as mean ± SD and categorical variables as counts (percentages). P-values reflect comparisons between genetically positive and genetically negative patients. RBD = REM sleep behavior disorder; MoCA = Montreal Cognitive Assessment; H&Y = Hoehn & Yahr stage

Genetic data was obtained from peripheral-blood DNA using three platforms available at different stages of recruitment: the NeuroBooster Array (NBA) [16], with quality control, genotyping calling and global ancestry as described in a LARGE-PD study [17]; a PD gene panel performed as part of the PD GENEration study [3]; and the TruSight One sequencing panel [18, 19] following previously published methods [20]. Platform use reflected test availability rather than a predefined selection strategy. To show the proportion of participants tested on each platform, the full cohort was used as the denominator; because some participants were tested on more than one platform during the study, the number of tests exceeded the number of participants (Supplementary Table 1). Local ancestry at the *LRRK2* locus was performed following a Global Parkinson’s Genetics program (GP2) pipeline [21].

Overall, the main PD-associated genes (*LRRK2, GBA1, PRKN, SNCA, PINK1, PARK7*, and *VPS35*) were assessed in all patients. Copy number variant (CNV) analysis and complete *GBA1* sequencing were available only for participants tested through the PD GENEration panel. For genes other than *GBA1*, variants classified as pathogenic or likely pathogenic according to the American College of Medical Genetics and Genomics (ACMG) guidelines [22] were considered as disease-causing. Monoallelic variants in autosomal recessive genes and variants of unknown significance were not included. As *GBA1* is considered a PD risk factor rather than a monogenic cause of disease [23], all *GBA1* variants were included irrespective of ACMG classification. Participants with genetic findings meeting these criteria—namely, a pathogenic or likely pathogenic variant in an autosomal dominant gene, biallelic pathogenic or likely pathogenic variants in an autosomal recessive gene, or *GBA1* variant—were considered as genetically positive (“positive”) for subsequent analyses; All other participants were classified as “negative”. When the same variant was detected in an individual using more than one testing platform, it was counted only once. in subsequent analyses.

All statistical analyses were performed in Python using relevant statistical libraries. Continuous variables were summarized as mean ± standard deviation, and categorical variables were summarized as absolute and relative frequencies.

## RESULTS

After excluding samples that failed quality control, the final cohort comprised 461 patients with PD. Overall, 57.3% of participants were tested using the NeuroBooster Array, 52.1% using the PD GENEration panel, and 21.7% using the TruSight One panel (Supplementary table 1). The mean AAO was 55.5 ± 12.2 years; 33.4% had early-onset PD (≤50 years), and 33.8% reported a positive FH. Positive participants more frequently reported a positive FH of PD (53.4% vs. 31.0%; p = 0.001) and had a higher prevalence of levodopa-induced dyskinesias (LID) (37.9% vs. 24.6%; p = 0.038). There were no other significant differences among groups (Table 1).

The genetic yield was 12.6%, with disease-associated variants identified in 58 of 461 participants. Variants were detected in *LRRK2, GBA1, PRKN, SNCA*, and *SQSTM1*, whereas no pathogenic or likely pathogenic variants were identified in *PINK1, PARK7*, or *VPS35* (Figure 1A). Among the positive cases, 29 carried the *LRRK2* p.G2019S variant, and 26 *GBA1* variants: (i) p.N370S (n = 13.8%), (ii) p.E365K (n = 6.9%), (iii) p.R496H (n =6.9%), (iv) p.L444P (n =5.2%), (v) p.T369M (n =5.2%), (vi) p.V457D (n=1), (vii) p.N188S (n=1), (viii) p.Leu422ProfsTer3 (n=1), (ix) p.Leu29AlafsTer18 (n=1). The remaining three participants carried a homozygous deletion of *PRKN* exons 8–9, a *SNCA* duplication, and a *SQSTM1* p.Pro392Leu variant.Genetic variants identified across testing platforms can be found in Supplementary Table 3.

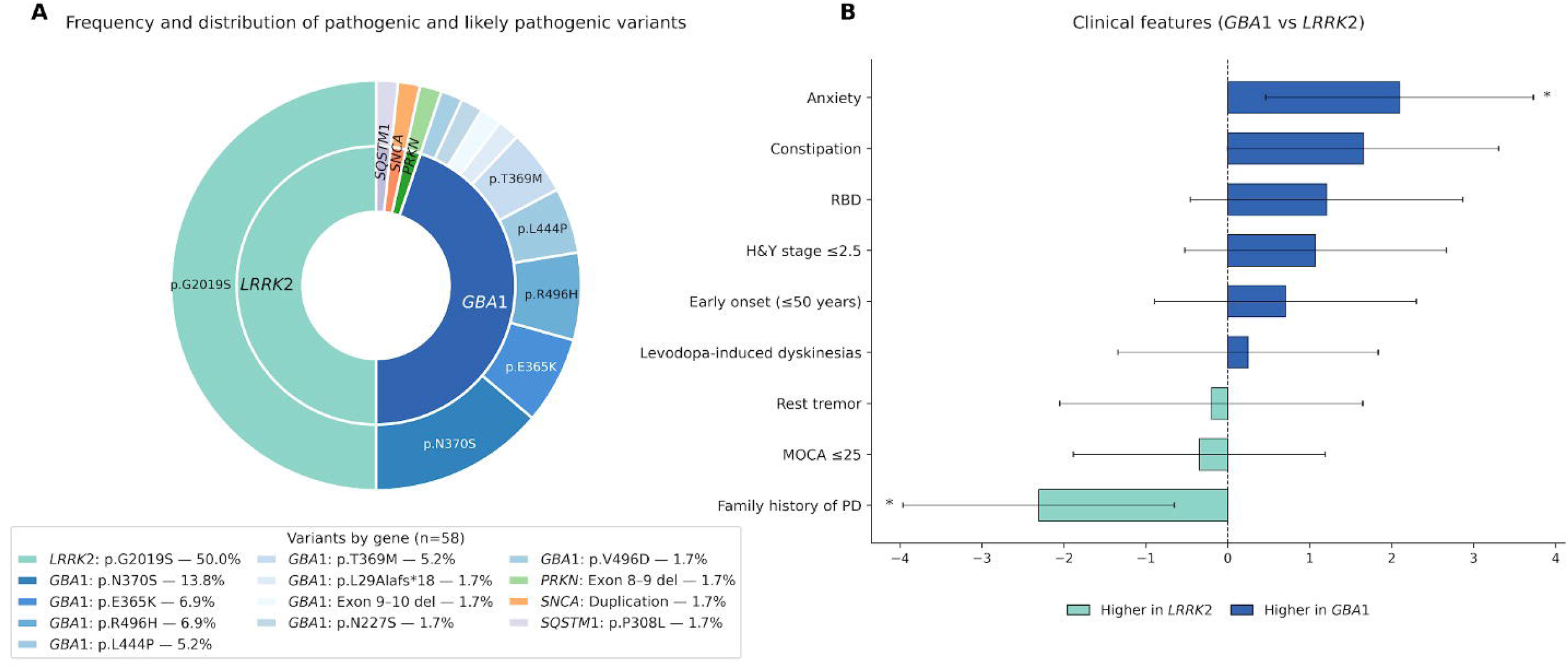

Clinical characteristics of genetically positive participants, including comparisons by gene, are summarized in Supplementary Table 2. A higher frequency of anxiety (69.2% vs. 34.5%; OR = 4.28, 95% CI 1.38–13.25; p = 0.015) was seen in *GBA1* carriers, while more frequent positive FH of PD among *LRRK2* carriers (72.4% vs. 34.6%; OR = 4.96, 95% CI 1.57–15.61; p = 0.007). A trend to younger AAO was seen in *GBA1* carriers, although not statistically significant (51.9 ± 10.1 vs. 57.1 ± 10.5 years; p = 0.071) (Supplementary Table 2 and Figure 1B).

The three participants with disease-associated variants in *PRKN, SNCA*, or *SQSTM1* showed heterogeneous clinical phenotypes. The participant with the homozygous *PRKN* exon 8–9 deletion had the earliest AAO (26-30 years), and was the only with a positive FH. The *SQSTM1* carrier developed PD between age 35-39 and had a positive response to deep-brain stimulation for motor complication. The whole-gene *SNCA* duplication participant had the latest AAO (51-55 years) and scored 23 on his MoCA score in the first year of disease.

Global ancestry showed that our cohort is a two-way admixed population (Supplementary Figure 1), with high contributions from Europeans (61.1%) and Native-Americans (37.8%). African, East Asian and South Asian ancestries combined represented 1.1 %, consistent with previous studies [24].

Local ancestry at the *LRRK2* locus was analyzed in 28 haplotypes from 14 *LRRK2* p.G2019S carriers and 522 haplotypes from 261 non-carriers. Ashkenazi Jewish ancestry was present in 11 of 28 carrier haplotypes (39.3%), compared with 23 of 522 non-carrier haplotypes (4.4%), indicating significant enrichment among *LRRK2* p.G2019S carriers (OR = 14.0, 95% CI 5.9–33.4; Fisher’s exact p = 9.8 × 10^-8^) (Supplementary Table 5).

## DISCUSSION

This study provides the largest clinical and genetic characterization of PD in Chile to date. Disease-associated variants were identified in 12.6% of patients, similar to recently reported in international cohorts [3, 4].

One of the main findings was the high frequency of *LRRK2* p.G2019S (6.3%) in this predominantly sporadic cohort, exceeding those previously reported for Chile and other South American cohorts [11, 12]. Its marked enrichment on Ashkenazi Jewish local ancestry supports a founder effect [25, 26], indicating a substantial Ashkenazi Jewish contribution to PD genetics in this cohort and illustrating the value of local ancestry analysis in admixed populations.

The *GBA1* variant spectrum may also reflect an Ashkenazi Jewish contribution, as p.N370S and p.R496H, both being more frequent in Ashkenazi Jewish populations, were among the most frequent variants identified [27, 28]. Although the overall *GBA1* frequency (5.6%) was consistent with previous Latin American and recent multi-ancestry reports [11, 29], the identification of nine distinct variants among 26 carriers highlights the ancestry-related heterogeneity of the Chilean population..

An unexpected finding was the identification of the *SQSTM1* p.Pro392Leu variant in one patient, as it has rarely been reported in PD, and it is more frequently reported in frontotemporal dementia and amyotrophic lateral sclerosis[30–32]. Its contribution to PD remains uncertain and needs further investigation in larger cohorts.

Clinical characteristics were largely similar between variant-positive and variant-negative patients, although variant carriers more frequently had a family history of PD and levodopa-induced dyskinesias (LID). The higher frequency of LID is consistent with Sosero et al.l [33]., who reported an increased risk of LID among *GBA1* carriers and a shorter time to LID onset on *LRRK2* carriers, suggesting gene-specific effects on levodopa-related motor complications. Among genetically defined subgroups, anxiety was more frequent in *GBA1* than in *LRRK2* carriers. This is consistent with reported phenotypic differences between genetic forms of PD, including a greater burden of psychiatric and other non-motor symptoms in *GBA1*-associated PD and a higher frequency of family history in *LRRK2*-associated PD [26–28, 34].

An important limitation of this study was the use of different testing platforms according to their availability over time, resulting in nonuniform analysis of genes and variants across the cohort. In particular, CNV analysis was available for only 52.1% of participants; therefore, deletions, duplications, and other structural variants involving genes such as *PRKN, PINK1*, and *SNCA* may have been missed. This may partly explain the low frequency of *PRKN* cases (0.2%). Nevertheless, this platform heterogeneity reflects the real-world constraints of conducting genetic research in resource-limited settings, particularly in low- and middle-income countries, where access to genetic testing remains limited[35]. Future studies incorporating systematic CNV analysis, and more uniform testing approaches may provide a fuller characterization of the genetic architecture of PD in the Chilean population.

In conclusion, this study characterizes the genetic landscape of PD in Chile. The high frequency of the *LRRK2* p.G2019S variant, together with the enrichment of Ashkenazi Jewish ancestry at this locus, supports an ancestry-related contribution to the frequency of this variant. The heterogeneous spectrum of *GBA1* variants further reflects the distinctive genetic architecture of PD in this admixed population. Together, these findings indicate that genetic patterns derived from predominantly European-ancestry cohorts may not fully represent admixed populations and reinforce the need for more inclusive research to advance equitable precision medicine.

## Ethics Statement

This study was approved by local Institutional Research Ethics Committee (“Comité de Ética de la Investigación del Servicio de Salud Metropolitano Norte”, approval number 017/2021) and conducted in accordance with the Declaration of Helsinki.

## Data Availability

The data supporting the findings of this study were generated as part of the LARGE-PD project. Individual-level data are not publicly available to protect participant privacy and confidentiality but may be accessed through LARGE-PD’s established data-access procedures, subject to the relevant approvals.

## Acknowledgment

We acknowledge the PD GENEration program (Parkinson’s Foundation) for providing genetic testing and genetic counseling for a subset of participants included in this study.

This work made use of the High-Performance Computing Cluster provided by the Cleveland Clinic Research Computing Services HPC (and other Linux-based analytic resources, such as the large, stand-alone R, Python and GPU/CUDA servers) are supported by the Cleveland Clinic Research Computing Services. Our work would not have been possible without the support of Eldon Walker, Ph.D., Director, LRI Computing Services and Michael Weiner, Senior HPC Administrator.

---

**Supplementary Table 1.a.** Platforms used in the cohort.

| Testing platform | N cases (% cohort) | Reported genes |
| --- | --- | --- |
| NeuroBooster Array | 264 (57.3 %) | *LRRK2, GBA1, PRKN, SNCA, PINK1, PARK7, VPS35* |
| PD GENeration panel | 240 (52.1 %) | *LRRK2, GBA1, PRKN, SNCA, PINK1, PARK7, VPS35* |
| TruSight One panel | 100 (21.7%) | *LRRK2, GBA1, PRKN, SNCA, PINK1, PARK7, VPS35, AFG3L2, ATP6AP2, ATP7B, DCAF17, NPC1, NPC2, NR4A2, OPA1, PDGFB, POLG, SCP2, SLC25A4, SLC25A46, SMPD1, SPG7, SQSTM1, TBK1, TWNK, VAC14, VCP, EPM2A, FA2H, ZFYVE26, ATP13A2, ATP1A3, C19orf12, CP, CSF1R, DCTN1, DNAJC6, FBXO7, FTL, GCH1, GLB1, GRN, LYST, MAPT, OPA3, PANK2, PDE8B, PDGFRB, PLA2G6, PRKRA, PTS, QDPR, RAB39B, SLC20A2, SLC30A10, SLC6A3, SPG11, SPR, TAF1, TH, TUBB4A, VPS13A, VPS13C, WDR45* |

Number and percentage of patients assessed with each genetic testing modality and genes reported for each platform. Percentages were calculated using the full cohort as the denominator (n = 461). Because some patients underwent more than one genetic test, testing modalities were not mutually exclusive and the total number of tests exceeds the number of patients.

**Supplementary Table 2.** Clinical and demographic features for all positive cases and stratified by *LRRK2* and *GBA1* carrier status.

| Variable | All positive (n=58) | *LRRK2* (n=29) | *GBA1* (n=26) | p-value | OR *GBA1* vs *LRRK2* (95% CI) | OR *LRRK2* vs *GBA1* (95% CI) |
| --- | --- | --- | --- | --- | --- | --- |
| Age at baseline | 63.40 ± 11.78 | 65.66 ± 10.95 | 59.54 ± 8.20 | **0.022** |  |  |
| Age of onset | 55.51 ± 12.17 | 57.07 ± 10.47 | 51.96 ± 10.10 | 0.071 |  |  |
| Time to diagnosis (years) | 1.54 ± 2.46 | 1.55 ± 2.64 | 1.62 ± 2.93 | 0.933 |  |  |
| Education (years) | 13.56 ± 4.12 | 13.69 ± 3.74 | 14.96 ± 2.42 | 0.137 |  |  |
| Disease duration (years) | 7.96 ± 6.23 | 8.59 ± 6.38 | 7.58 ± 5.14 | 0.519 |  |  |
| Early onset (≤50 years) | 154 (33.4%) | 9 (31.03%) | 11 (42.3%) | 0.415 | 1.63 (0.54–4.93) | 0.61 (0.20–1.86) |
| Male | 272 (59.0%) | 17 (58.62%) | 15 (57.7%) | 1.000 | 0.96 (0.33–2.81) | 1.04 (0.36–3.04) |
| Family history | 156 (33.8%) | 21 (72.41%) | 9 (34.6%) | **0.007** | 0.20 (0.06–0.64) | 4.96 (1.57–15.61) |
| Rest tremor | 340 (73.8%) | 23 (79.31%) | 20 (76.9%) | 1.000 | 0.87 (0.24–3.13) | 1.15 (0.32–4.14) |
| Hyposmia | 207 (44.9%) | 14 (48.28%) | 16 (61.5%) | 0.419 | 1.71 (0.59–5.02) | 0.58 (0.20–1.71) |
| Constipation | 198 (43.0%) | 7 (24.14%) | 13 (50.0%) | 0.056 | 3.14 (1.00–9.89) | 0.32 (0.10–1.00) |
| Depression | 190 (41.2%) | 12 (41.38%) | 12 (46.2%) | 0.789 | 1.21 (0.42–3.53) | 0.82 (0.28–2.40) |
| Anxiety | 206 (44.7%) | 10 (34.48%) | 18 (69.23%) | **0.015** | 4.28 (1.38–13.25) | 0.23 (0.08–0.73) |
| Sleep problems | 227 (49.2%) | 16 (55.17%) | 17 (65.38%) | 0.583 | 1.53 (0.52–4.57) | 0.65 (0.22–1.94) |
| RBD | 171 (37.1%) | 7 (24.14%) | 11 (42.31%) | 0.249 | 2.30 (0.73–7.30) | 0.43 (0.14–1.37) |
| Levodopa response | 352 (76.4%) | 20 (68.97%) | 21 (80.77%) | 0.367 | 1.89 (0.54–6.62) | 0.53 (0.15–1.85) |
| Levodopa-induced dyskinesias | 121 (26.2%) | 10 (34.48%) | 10 (38.46%) | 0.786 | 1.19 (0.40–3.57) | 0.84 (0.28–2.53) |
| MOCA ≤25 | 268 (58.1%) | 14 (48.28%) | 11 (42.31%) | 0.788 | 0.79 (0.27–2.28) | 1.27 (0.44–3.69) |
| H&Y stage ≤2.5 | 331 (71.8%) | 15 (51.72%) | 18 (69.23%) | 0.271 | 2.10 (0.69–6.35) | 0.48 (0.16–1.44) |

Mean ± SD for continuous traits; counts (%) for categorical traits. Welch's t-test was used for continuous variables and Fisher's exact test for categorical comparisons. Bold values indicate nominal statistical significance ( $p < 0.05$ ). OR = odds ratio; CI = confidence interval; RBD = REM sleep behavior disorder; MOCA = Montreal Cognitive Assessment; H&Y = Hoehn & Yahr stage.

**Supplementary Table 3.** Genetic variants identified across testing platforms.

| Gene | n | Variant | NeuroBooster Array | TruSight One panel | PD GENERation panel |
| --- | --- | --- | --- | --- | --- |
| *LRRK2* | 29 | p.Gly2019Ser (G2019S) | ✓ | ✓ | ✓ |
| *GBA1* | 8 | p.Asn409Ser (N370S) | ✓ | ✓ | ✓ |
| *GBA1* | 4 | p.Glu365Lys (E326K) | ✓ | — | ✓ |
| *GBA1* | 4 | p.Arg535His (R496H) | ✓ | — | ✓ |
| *GBA1* | 3 | p.Thr408Met (T369M) | ✓ | ✓ | — |
| *GBA1* | 3 | p.Leu483Pro (L444P) | — | — | ✓ |
| *GBA1* | 1 | p.Val496Asp (V457D) | ✓ | — | — |
| *GBA1* | 1 | p.Asn227Ser (N188S) | ✓ | ✓ | — |
| *GBA1* | 1 | p.Leu422ProfsTer3 (1263del55) | — | — | ✓ |
| *GBA1* | 1 | p.Leu29AlafsTer18 (L(-11)Afs*18) | — | — | ✓ |
| *SNCA* | 1 | Whole-gene duplication | — | — | ✓ |
| *PRKN* | 1 | Homozygous deletion of exons 8–9 | — | — | ✓ |
| *SQSTM1* | 1 | p.Pro392Leu (P392L) | — | ✓ | — |

Checkmarks indicate that the variant was identified in at least one participant using the corresponding testing platform. Some participants underwent testing with more than one platform.

**Supplementary Table 4.** Clinical characteristics of patients with disease-causing genetic variants.

| Gene | Variant | Gender | AAO | DD | H&Y | FH | Non-motor symptoms | LID | Others | MOCA test |
| --- | --- | --- | --- | --- | --- | --- | --- | --- | --- | --- |
| LRRK2 | p.Gly2019Ser | Female | 71-75 | 10 | 4 | No | Hyposmia, anxiety, insomnia, RBD | No | - | 27 |
|  | p.Gly2019Ser | Female | 46-50 | 16 | 2 | Yes, second degree | Hyposmia, anxiety, depression, insomnia | Yes | Postural instability | 25 |
|  | p.Gly2019Ser | Male | 56-60 | 3 | 2 | Yes, second degree | Insomnia | No | - | 27 |
|  | p.Gly2019Ser | Male | 66-70 | 3 | 3 | Yes, first degree | No | No | <70% levodopa response | 30 |
|  | p.Gly2019Ser | Female | 61-65 | 17 | 3 | Yes, first degree | Hyposmia, constipation, anxiety, depression, insomnia | Yes | No rest tremor, Postural instability | 20 |
|  | p.Gly2019Ser | Female | 56-60 | 23 | 3 | Yes, first degree | Insomnia | Yes | Postural instability | 19 |
|  | p.Gly2019Ser | Male | 56-60 | 13 | 1 | Yes, first degree | Hyposmia, insomnia, RBD | Yes | Postural instability | 27 |
|  | p.Gly2019Ser | Female | 66-70 | 2 | 3 | No | Insomnia | No | - | 26 |
|  | p.Gly2019Ser | Male | 66-70 | 3 | 2 | Yes, first degree | Anxiety, depression | Yes | Postural instability | 8 |
|  | p.Gly2019Ser | Female | 36-40 | 7 | 3 | No | No | No | Postural instability | 25 |
|  | p.Gly2019Ser | Male | 46-50 | 22 | 3 | Yes, first degree | Depression, insomnia | No | Postural instability | 26 |
|  | p.Gly2019Ser | Male | 36-40 | 12 | 1 | No | Hyposmia, depression, RBD | No | No rest tremor, Postural instability | 23 |
|  | p.Gly2019Ser | Male | 46-50 | 5 | 2 | Yes, first degree | Hyposmia, constipation, anxiety, depression, RBD | No | Postural instability, <70% levodopa response | 27 |
|  | p.Gly2019Ser | Male | 46-50 | 10 | 1,5 | Yes, first degree | Insomnia | Yes | No rigidity | 30 |
|  | p.Gly2019Ser | Male | 66-70 | 0 | 2 | Yes, first degree | Hyposmia, anxiety, depression, insomnia | No | - | 19 |
|  | p.Gly2019Ser | Female | 61-65 | 7 | 1,5 | Yes, first degree | Anxiety, depression, insomnia, RBD | No | - | 22 |
|  | p.Gly2019Ser | Male | 41-45 | 3 | 2 | No | Insomnia | No | No rest tremor | 28 |
|  | p.Gly2019Ser | Male | 56-60 | 12 | 3 | Yes, first degree | Insomnia | Yes | - | 26 |
|  | p.Gly2019Ser | Female | 66-70 | 5 | 2 | No | Hyposmia, constipation, depression, insomnia | Yes | Postural instability | 21 |
|  | p.Gly2019Ser | Female | 56-60 | 18 | 2 | Yes, first degree | Hyposmia, insomnia | No | Postural instability | 21 |
|  | p.Gly2019Ser | Male | 51-55 | 2 | 2 | Yes, second degree | No | No | No rest tremor | 25 |
|  | p.Gly2019Ser | Male | 46-50 | 2 | 3 | Yes, second degree | Anxiety | No | - | 25 |
|  | p.Gly2019Ser | Male | 56-60 | 1 | 1 | Yes, first degree | Hyposmia, depression | No | No rest tremor, Postural instability | 28 |
|  | p.Gly2019Ser | Male | 61-65 | 3 | 4 | Yes, first degree | Hyposmia, constipation, RBD | No | No rigidity, <70% levodopa response | 27 |
|  | p.Gly2019Ser | Female | 76-80 | 7 | 4 | Yes, first degree | Hyposmia, constipation | No | Postural instability | 5 |
|  | p.Gly2019Ser | Male | 36-40 | 14 | 4 | Yes, second degree | Hyposmia, anxiety, depression, insomnia, RBD | No | Postural instability | 26 |
|  | p.Gly2019Ser | Male | 51-55 | 8 | 3 | No | Hyposmia, anxiety | Yes | Postural instability | 26 |
|  | p.Gly2019Ser | Female | 66-70 | 9 | 2,5 | No | Constipation | No | No rigidity, Postural instability, <70% levodopa response | 29 |
|  | p.Gly2019Ser | Female | 51-55 | 12 | NA | Yes, first degree | Constipation, depression | Yes | Postural instability | 22 |
| GBA1 | p.Asn409Ser (N370S) | Female | 66-70 | 8 | 3 | Yes, first degree | Anxiety, RBD | No | No rest tremor, postural instability | 21 |
|  | p.Asn409Ser (N370S) | Female | 61-65 | 5 | 2 | No | Hyposmia, constipation, anxiety, depression, insomnia, RBD | No | - | 30 |
|  | p.Asn409Ser (N370S) | Male | 51-55 | 7 | 2 | No | Hyposmia | No | - | 26 |
|  | p.Asn409Ser (N370S) | Male | 36-40 | 11 | 3 | No | Hyposmia, anxiety, depression | Yes | Postural instability | 27 |
|  | p.Asn409Ser (N370S) | Male | 51-55 | 10 | 3 | No | Hyposmia, constipation, anxiety, depression, insomnia, RBD | Yes | No rest tremor, postural instability | 20 |
|  | p.Asn409Ser (N370S) | Female | 36-40 | 10 | 2 | No | Constipation, anxiety, depression, insomnia | Yes | - | 19 |
|  | p.Asn409Ser (N370S) | Male | 51-55 | 5 | 2 | No | Anxiety, depression, insomnia, RBD | No | - | 24 |
|  | p.Asn409Ser (N370S) | Male | 56-60 | 1 | 1 | No | Hyposmia, constipation, anxiety, insomnia | No | - | 28 |
|  | p.Glu365Lys (E326K) | Male | 36-40 | 9 | 1,5 | No | Hyposmia, insomnia, RBD | Yes | <70% levodopa response | 27 |
|  | p.Glu365Lys (E326K) | Male | 31-35 | 8 | 2,5 | No | Constipation, insomnia | No | Postural instability | 27 |
|  | p.Glu365Lys (E326K) | Female | 61-65 | 6 | 2 | No | Hyposmia, insomnia | No | - | 27 |
|  | p.Glu365Lys (E326K) | Male | 41-45 | 22 | 2 | No | Constipation | Yes | No rest tremor | 24 |
|  | p.Arg535His (R496H) | Male | 66-70 | 3 | 1,5 | No | Constipation, anxiety, depression, insomnia, RBD | No | No rest tremor | 24 |
|  | p.Arg535His (R496H) | Female | 36-40 | 16 | 4 | Yes, first degree | Hyposmia, constipation, anxiety, depression, insomnia, RBD | Yes | No rest tremor, postural instability | 29 |
|  | p.Arg535His (R496H) | Male | 61-65 | 2 | 1 | Yes, second degree | Hyposmia, anxiety | No | - | 28 |
|  | p.Arg535His (R496H) | Male | 36-40 | 15 | 4 | No | Hyposmia, anxiety, RBD | Yes | Postural instability | NA |
|  | p.Leu483Pro (L444P) | Male | 51-55 | 2 | 1,5 | No | Constipation, anxiety, insomnia | No | Postural instability | 22 |
|  | p.Leu483Pro (L444P) | Male | 51-55 | 5 | 2 | No | Hyposmia, insomnia, RBD | Yes | - | 22 |
|  | p.Leu483Pro (L444P) | Male | 46-50 | 8 | 3 | Yes, first degree | Hyposmia, anxiety, depression | Yes | Postural instability | 10 |
|  | p.Thr408Met (T369M) | Female | 61-65 | 5 | 1 | No | Insomnia, RBD | No | - | 27 |
|  | p.Thr408Met (T369M) | Female | 46-50 | 9 | 2,5 | No | Hyposmia, anxiety, depression, insomnia | No | Postural instability, <70% levodopa response | 25 |
|  | p.Thr408Met (T369M) | Female | 46-50 | 5 | 3 | No | Anxiety, depression | No | - | 26 |
|  | p.Asn227Ser (N188S) | Female | 56-60 | 3 | 3 | Yes, second degree | Hyposmia, constipation, anxiety, depression, insomnia | No | - | 19 |
|  | p.Val496Asp (V457D) | Female | 61-65 | 4 | 2 | Yes, first degree | Hyposmia, constipation, anxiety, insomnia | Yes | Postural instability | 26 |
|  | p.Leu422ProfsTer3 (1263del55) | Male | 51-55 | 2 | 1 | Yes, first degree | Hyposmia, constipation, RBD | No | Postural instability | 27 |
|  | p.Leu29AlafsTer18 (L(-11)Afs*18) | Female | 46-50 | 16 | 2 | Yes, second degree | Constipation, anxiety, depression, insomnia | No | - | 29 |
| PRKN | Homozygous Deletion of Exons 8-9 | Female | 26-30 | 16 | 2,5 | Yes, first degree | Hyposmia, anxiety, depression, RBD | Yes | No rest tremor | 25 |
| SQSTM1 | p.Pro392Leu (P392L) | Male | 31-35 | 10 | 2 | No | Constipation, anxiety, depression, insomnia | Yes | Early falls, deep-brain stimulation with a positive result | 19 |
| SNCA | Whole Gene Duplication | Male | 51-55 | 1 | 1,5 | No | Depression, insomnia, RBD | No | - | 23 |

AAO: range of age of onset, DD: disease duration, FH: presence of family member with Parkinson's Disease, RBD: REM sleep behavior disorder, LID: presence of severe levodopa induced dyskinesia, H&Y: Hoehn and Yahr scale. Ancestry and Ethnicity are self reported.

**Supplementary Table 5.** Local ancestry at the *LRRK2* G2019S locus.

| Group | AJ haplotypes, n (%) | Non-AJ haplotypes, n (%) | Total haplotypes | OR | 95% CI | p-value |
| --- | --- | --- | --- | --- | --- | --- |
| LRRK2 p.G2019S carriers | 11 (39.3) | 17 (60.7) | 28 | 14.0 | 5.9–33.4 | $9.8 \times 10^{-8}$ |
| Non-carriers | 23 (4.4) | 499 (95.6) | 522 | Reference | — | — |

AJ, Ashkenazi Jewish; CI, confidence interval; OR, odds ratio. Local ancestry was evaluated at the *LRRK2* G2019S locus and summarized at the haplotype level. Percentages were calculated within each carrier-status group. The odds ratio, 95% confidence interval, and p-value were estimated using Fisher's exact test comparing AJ versus non-AJ haplotypes in *LRRK2* p.G2019S carriers and non-carriers.

**Supplementary Figure 1:**
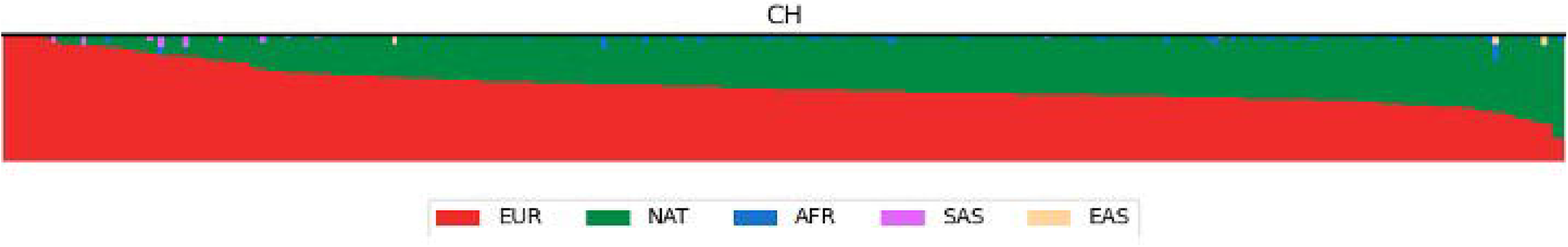
Global ancestry analysis of the cohort. Each vertical bar represents one participant and shows the estimated proportions of European (EUR, red), Native American (NAT, green), African (AFR, blue), South Asian (SAS, pink), and East Asian (EAS, yellow) ancestry.

